# Covid-19 respiratory protection: the filtration efficiency assessment of decontaminated FFP2 masks responding to associated shortages

**DOI:** 10.1101/2021.01.18.21249976

**Authors:** M. Benboubker, B. Oumokhtar, F. Hmami, K. El Mabrouk, L.EL Alami, B. Arhoune, M F. Belahsen, A. Aboutajeddine

## Abstract

During the Covid-19 pandemic, healthcare workers were extremely vulnerable to infection with the virus and needed continuous protection. One of the most effective and widely used means of protection was the FFP2 respirator. Unfortunately, this crisis created a shortage of these masks, prompting hospitals to explore opportunities to reuse them after decontamination.

An approach for assessing the filtration efficiency of decontaminated FFP2 masks has been proposed and applied to evaluate the possibilities of their safe reuse. The decontamination processes adopted are those based on moist heat or hydrogen peroxide. The approach introduces efficiency measures that define the filtration and protection capacity of the masks, which characterize both chemical and structural changes, and encompasses many techniques including scanning electron microscopy (SEM), Fourier transforms infrared spectroscopy (FTIR), and thermogravimetric analysis (TGA). The test protocol was applied to mask samples that had endured different decontamination cycles and the results of their efficiency measures were compared to brand-new masks’ performances.

The main result was that chemical and structural characterization of the decontaminated masks have shown no substantial change or deformation of their filter media structures. Indeed, the respiratory resistance test has shown that the results of both the FFP2 masks that have undergone a hydrogen peroxide disinfection cycle or a steam autoclave cycle remained constant with a small variation of 10 Pa from the EN149 standard. The chemical characterization, on the other hand, has shown that the filter media of the decontaminated masks remains unchanged, with no detectable chemical derivatives in its constituents.

## Introduction

The COVID-19 pandemic has triggered an acute issue of ensuring the distribution and deployment of personal protective equipment (PPE) for healthcare workers to manage patients. This situation had led the World Health Organization (WHO) to warn on the increased demand for personal protective equipment (*PPE*) and their possible shortages. In effect, the fast-growing flow of infected patients and the worldwide impact of the outbreak has strained supply chains, not only for equipment such as ventilators but also for disposable PPE used to manage patients, including disposable respirator masks (FFP2).

FFP2 masks are an essential element of personal protective equipment used in healthcare settings ^1 2 3 4^. These masks can filter particles with a size ranging from 0.4 to 0.6 μm with an efficiency of 94%. Generally, they are the most recommended for respiratory precautions ^2^. Currently, these masks are intended for single-use due to potential microbial and viral contamination. However, the recent Covid-19 pandemic has resulted in a severe shortage of these masks, which prompted the study of various decontamination procedures for possible reuse. The evaluation of the potential reuse of masks is therefore a rapidly emerging necessity to address the problem of this shortage and to ensure an optimal protection of healthcare workers.

Many attempts have been documented to propose solutions around the world to solve the FFP2 mask shortage and to provide reassuring arguments for possible safe and secure reuse. Several processes have been reported, including dry and wet heat, hydrogen peroxide vapor, ozone, UV radiation, etc. These studies undertaken in this area have proposed an approach to evaluate the filtration performance of respiratory masks after decontamination according to the standards^5 6 7^. However, there is a suspicion that these processes alter the filtration capacity of these masks, mainly those using hydrogen peroxide vapors ^8^.

On the other hand, the proposal of a reuse protocol depends on the availability of a reprocessing method that should be simple, safe, and adapted to the hospital context, which includes the logistic organization and the human factor. These requirements, particularly availability and simple supply chain scenarios in the Moroccan hospitals, have led to the choice of reprocessing methods based on the moist heat and hydrogen peroxide vapors, which were also the most methods used during this Covi-19 pandemic context. The goal of this work is to examine possible structural and chemical changes of the FFP2 masks after exposure to the chosen reprocessing methods. Explicitly, to control environmental risks, we tested (only) brand new FFP2 of the brand Duckbill, which are the most used in Moroccan hospitals, after reprocessing cycles up to four times by each method. The purpose of this protocol is to study the filtering and mechanical properties of the decontaminated FFP2 masks.

## Material and method

### Filtering mechanisms

The FFP2 masks can filter foreign particles by intercepting them in different material layers of the mask, thanks to physical phenomena that can be mechanical or electrostatic ^9^. The particles usually pass through the mask in a laminar flow, so that the flow would generally bend smoothly around obstacles (fibers). If this is the case, the mechanical capture of the particles at the surface of the fibers occurs either, when the inertia of the particle is large enough to deviate from its aerodynamic trajectory, or the suspended particles do not follow the air streamlines but meet the air molecules and take a disordered and random trajectory. This vibratory motion increases the probability that the particles get into touch with the fibers and remain suspended in them due to Van der Waals’s weak molecular interaction. Generally, this mechanism helps trap airborne particles with a diameter of around 0.1 μm ^10 11^.

Besides, a mechanism of electrostatic capture of particles (charged or not) is possible when the charges of the fibers are extremely important ^11^, Fibers can attract both intrinsically charged particles by Colombian forces and neutral polar particles (such as tiny water droplets) by dielectrophoresis forces resulting from the interaction of polarized objects and electric field gradients. Most FFP2 masks depend on both mechanical and electrostatic effects to achieve the filtration effect. In this sense, the evaluation of the filtration efficiency of this type of masks is based on several tests and parameters according to the standards in this field.

### Measurements of respiratory mask efficiency

Standards list different measurable parameters that can characterize the respiratory mask efficiency. The most measures, in similar studies, which aim at the reuse of respirators are the respiratory resistance, the aerosolization tests, and the leak tests. In this study, we have taken into consideration some parameters usually tested in the field of manufacturing for this type of filter. The measures considered in this study are:

#### Measure 1: Respiratory resistance

This parameter, taken into consideration by ISO EN 149 standard, includes two essential measures, the inhalation, and the breathing resistance. The test is performed for 3 minutes and indicates the values of efficiency and respiratory resistance by applying two flow rates, one simulating inspiration (95 l / min -0.2 m / s) and the second, expiration (160 l / min - 0.34 m / s).

#### Measure 2: The lifetime and respiratory comfort

This parameter is mainly related to the performance of tight-fitting respirators that are supposed to achieve a good seal between the facepiece of the respirator and the wearer’s face. If the seal is inadequate, contaminated air takes the path of least resistance and travels through leaks in the face seal. Consequently, a poor seal to the face unavoidably reduces the level of protection provided to the wearer. This can impact the lifetime of the product, estimated between four and eight hours ^12^.

#### Measure 3: The quality of distribution and the intermingling of fibers

The best-known manufacturing process for FFP2 masks’ non-woven fabrics is the melting process that gives fine fibers. The fineness of the fibers obtained is usually between 0,6 and 10 μm for the meltblown and between 1 to 50 μm for the spunbond. This parameter is essential to obtain a high filtering efficiency.

#### Measure 4: Chemical structure

The layers and the filter media of masks studied are generally based on Polypropylene. On the other hand, Exposure of Polypropylene to temperature causes deterioration of the material and loss of its mechanical properties. During its degradation, the formation of oxidation products such as hydro-peroxides and various carbonyl products are highlighted in this study. They can influence the filtering power of our masks which results in the appearance of derivatives of chemical compounds. In this study, we have proceeded to chemical characterization to explore the molecular changes of the filter media. It should be remembered that some common thermoplastics, such as polypropylene (PP), have such a low thermal degradation plateau^13 14 15^.

#### Measure 5: moisture saturation

Moisture accumulation reduces the filtering efficiency of FFP2 masks. Therefore, each type of mask has a maximum usage time that depends on the ability of its materials to absorb moisture ^16^. In this work, we have taken into consideration this parameter which can affect the filtration efficiency of our masks, especially as we use a decontamination protocol based on moist heat ^13 14^.

## Testing protocol

### FFP2 mask decontamination protocols

The masks were divided into two samples: the first was exposed to moist heat sterilization (121°C for 20 min, with 15 min drying time) and the second to hydrogen peroxide doped with silver ions (12% H2O2 at a rate of 6 ml per m3 for 10 minutes and a contact time of one hour on a 27 m3 enclosure), For this purpose, we have used a rotating nozzle micro-nebulizer to generate particles smaller than 5 µm to ensure smooth penetration.It is important to note that to ensure the sterilization performance validation, we have used multiparametric tests and Bowie’s dick penetration tests as required by international standards and guidelines (ISO 17665).

### Samples studied

The samples analyzed are FFP2 (Duckbill) type masks. The composition of the layers and the filter media is generally based on Polypropylene. After reprocessing the parts of the samples studied are the inner and outer layers (Spunbond) and the intermediate filter media (Melt-blown) of the mask. The protocol used takes into account a maximum of four reuses of the FFP2 mask. Indeed, the protocol consists of performing decontamination tests on twenty new masks to protect the manipulators during the study. This protocol also includes the study of a control sample of FFP2 masks without any exposure to any reprocessing procedure.

### Analysis methods

#### Respiratory resistance test

According to the test standard ISO EN 149,this test consists of measuring the resistance to airflow through the whole respirator at various flow rates designed to represent the peak inhalation and exhalation rates of a wearer during work of low to moderate intensity (including peak flow rates of 95lpm, the minute volume of 30lpm, and 160lpm, minute volume of 50lpm). The maximum permitted breathing resistance is 3.0mbar.To perform the test, we have used a motorized differential pressure sensor adapted to the test forces and standard accessories for fixing and holding FFP2 masks on the test device.

#### Scanning Electron Microscopy (SEM)

The morphological properties, quality of distribution, and the intermingling of fibers have been studied by Quanta 200 FEI equipped with EDX probe with software Genesis 2000i and 15 kV accelerating voltage. The samples were mounted on adhesive tape and a water-coated nozzle to improve electrical conductivity. The samples included in this type of analysis are the filter media (Melt-blown) and the external and internal layers (Spunbond) of the FFP2 masks that are the object of this study.

#### Fourier transforms infrared spectroscopy (FTIR)

The chemical structure changes of the reprocessed masks are characterized through the examination of the FTIR spectra. These spectra were obtained using Spectrometer FT- IR Nicolet iS50, piloted and recorded in the frequency range between 450 to 4000 cm^-1^ using a resolution of 4 cm^-1^ with a sampling frequency of 16 scans. The samples included in this type of analysis are the filter media of the masks (Melt-blown).

#### Thermogravimetric Analysis (TGA)

To observe possible moisture saturation in the layers of the FFP2 masks, the thermogravimetric behavior of our samples was studied. This was done on a Q500 TGA device. The samples of approximately 10 mg were heated from 25 to 500 °C at a heating rate of 10 °C/min under 20 ml/min of nitrogen current. The samples examined for this type of analysis are the filter media of the masks FFP2 (Melt-blown).

### Analysis and interpretation

All the above tests have been performed on decontaminated samples as well as the control sample. The data obtained were analyzed to study the influence of the decontamination process on the filtering efficiency and filter media material (Melt-blown, spunbond) of the FFP2 masks, using an empirical analytical approach.

## Results

### Respiratory resistance

To study the filtration efficiency of our masks after exposure to the reprocessing processes, respiratory resistance tests (inspiration and expiration) were conducted on our samples that underwent different decontamination cycles. The test includes a motorized differential pressure sensor adapted to the test forces and standard accessories for fixing and holding FFP2 masks on the test device. The results are reported in Table I.

**Table I.** Respiratory resistance results according to EN149 standard

| **Masks sample** | **Expiratory resistance (Pa)**<br>**160 l/min – 0,34 m/s** | **Inspiratory resistance (Pa)**<br>**95 l/min -0,2 m/s** |
| --- | --- | --- |
| *Witness FFP2 Masks\** | 340 | 350 |
| *H2O2/Ag<sup>+</sup> FFP2 Masks<sup>±</sup>* | 340 | 340 |
| *One steam cycle FFP2 Masks<sup>‡</sup>* | 360 | 320 |
| *Two steam cycle FFP2 Masks<sup>‡</sup>* | 330 | 330 |
| *Three steam cycle FFP2 Masks<sup>‡</sup>* | 330 | 330 |
| *Four steam cycle FFP2 Masks<sup>‡</sup>* | 330 | 330 |

\*: New masks FFP2, no decontamination was received
±: Masks FFP2, have been decontaminated with hydrogen peroxide and silver ions.
‡: Masks FFP2, received a moist heat decontamination.

These results show that the respiratory resistance of new FFP2 masks is 340 Pa at exhalation and 350 Pa at inspiration. This deviation from EN149, which is in the order of -/+ 50 Pa, maybe due to a storage defect or exposure to moisture or radiation^17 18^, as these types of masks are electret type filters, they have to be stored under conditions recommended by the manufacturer. The effects of mask storage have already been studied in the literature. It is reported that for a stock of more than 10 years old for some masks, 10% no longer fulfill the efficiency criterion and are below the standard ^19^. However, with this defect, the filtration efficiency is relatively high, or always above 90%.

For the FFP2 masks that underwent a single cycle of hydrogen peroxide and silver ion disinfection or a steam heat autoclave cycle, the performances of respiratory resistance have remained constant with a small difference of 10 Pa in comparison with the witness FFP2, a small difference of 0.02% at inspiration.

FFP2 masks that underwent two to four cycles of disinfection by steam autoclaving showed a decrease of the order of 20 Pa on inspiration and expiration compared to the witness FFP2 masks. In general, the uncertainty on this means of experimentation adopted in our study is +/- 0.01, which leads us to conclude that the proposed reprocessing techniques are acceptable. Indeed, these results obtained on the respiratory resistance test allow possible reuse of three FFP2 masks according to the EN149 standard ^20^.

### Scanning Electron Microscopy (SEM)

The quality of distribution and the intermingling of the fibers on the surface is evaluated. Sectional views are taken to observe the organization of the fibers on the thickness as well as the different fiber mixtures in the filtering media (Melt-blown) and the external layers of the FFP2 masks (Spunbond).

The magnifications range was from x200 to x400. The resolution of the images obtained is 2048 x 1536 pixels with always the pixel as the conversion unit for the measurement of the fiber diameter.

Many types of FFP2 respirator masks are currently commercialized, these are composed according to a common manufacturing process of two polypropylene Spunbond protective layers and two or more Melt-blown filtering layers, each of these layers will be studied separately.

The data concerning the thickness and diameter of the fibers are grouped in Table II and Figure I SEM. The different operating procedures explained are detailed in the methodology section. The aim here is to get an idea of the structural characteristics of the layers studied to report possible structural changes in the SEM of the exposed masks compared to the witness FFP2 mask.

**Table II.**
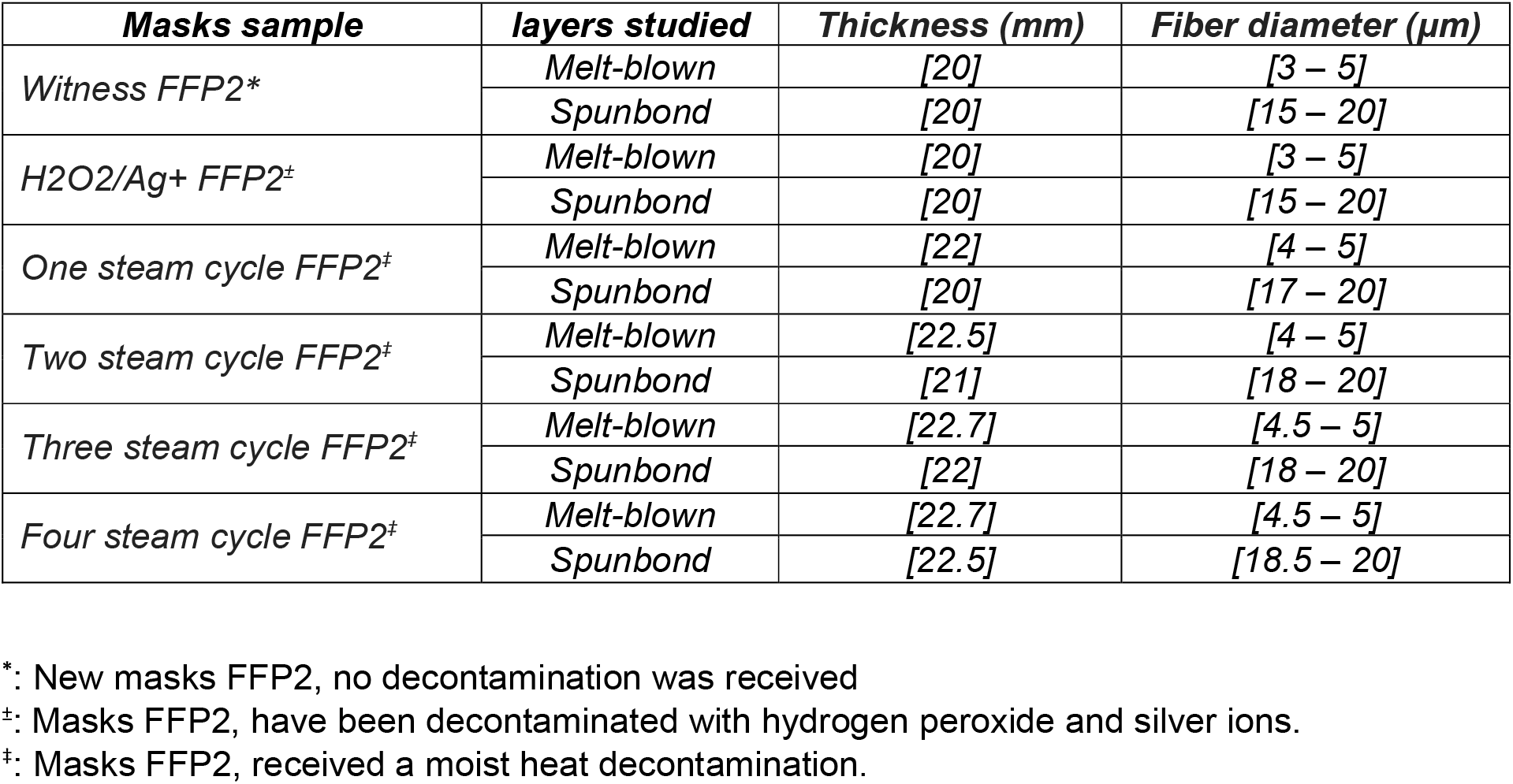
Structural characteristics measured on monolayers composing FFP2 masks per reprocessing cycle.

**Figure I.**
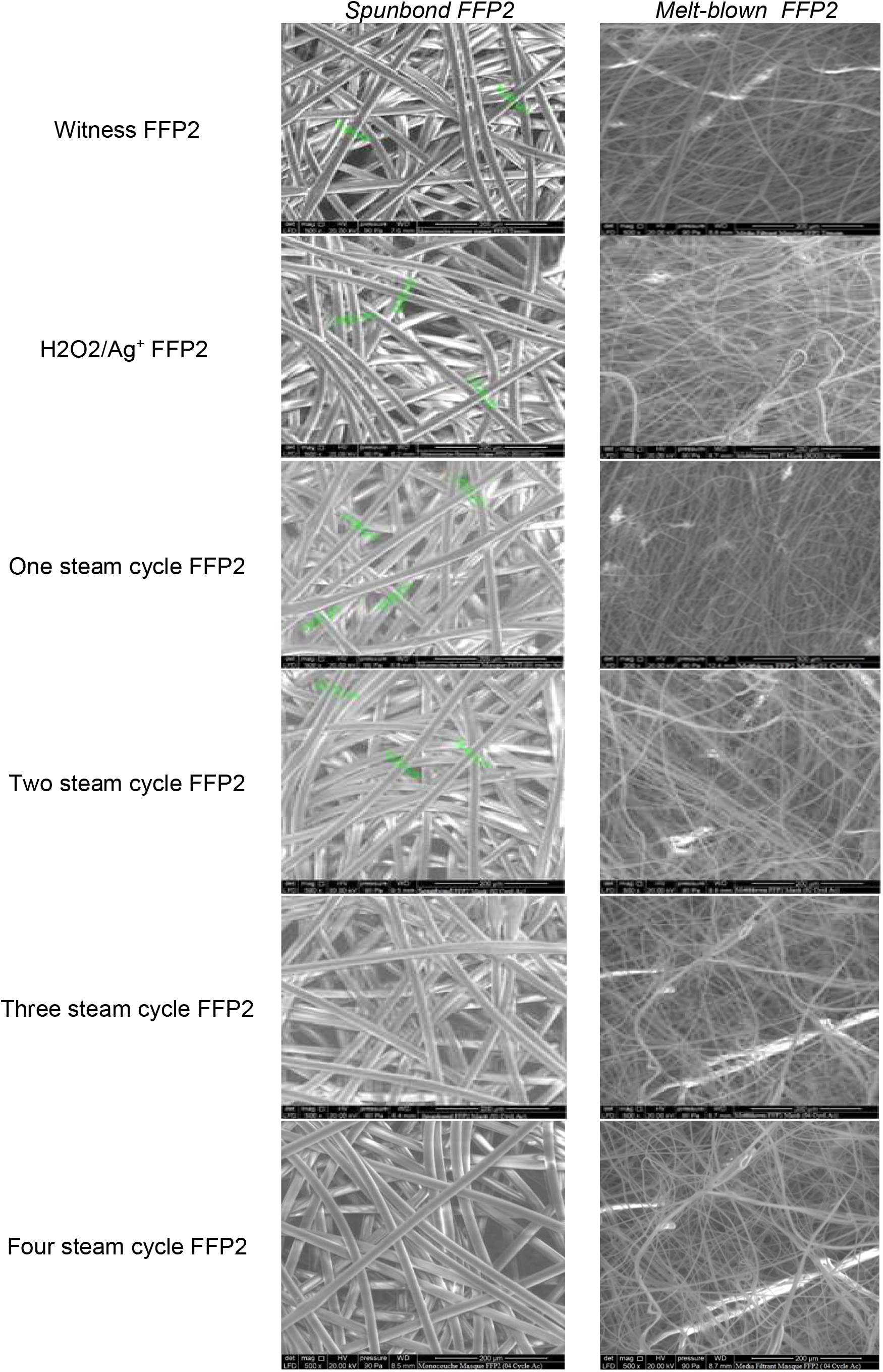
SEM images of the *Melt-blown* and *Spunbond composing FFP2 masks per reprocessing cycle*.

In the manufacturing process for FFP2 masks, the Melt-blown layers are less compact than those of Spunbond, the Melt-blown process makes it possible to obtain smaller fibers necessary for the filtration efficiency of FFP2 masks^21 22^. The objective of this part is to observe a possible modification of the fiber diameters, especially since the reprocessing method adopted is steam heat. The latter can also influence the compactness of the filter media which can be clogged due to the dryness during autoclaving of the FFP2 masks and subsequently to an increase in the pressure drop and the permeance of the Melt- blown layers ^23 24 25^.

Control of fiber diameter seems more difficult to judge from Figure I, which shows the fiber diameter distribution of the Melt-blown and Spunband samples of our FFP2 masks in this study. This distribution shows a great disparity concerning the diameter of the fibers in the same sample, especially for the filtering media, which is a characteristic of Melt- blown products in general ^24^.

The results obtained show that the diameter of the fibers in the samples treated with hydrogen peroxide and silver ions remains constant compared to the control samples, in contrast to the samples exposed to autoclaving by steam heat, which showed an increase in diameter from 0.5 to 3.5 μm in the spunbond layers and from 2 to 2.5 mm in thickness, which can be explained by the saturation of humidity in the samples studied (Figure I, Table II).

These results obtained may lead us to conclude that our masks have not changed much structurally, the parameters obtained from the respiratory resistance tests confirm this hypothesis since it has decreased especially for FFP2 masks undergoing steam heat processing cycles.

### Fourier transforms infrared spectroscopy (FTIR)

The IR absorption spectroscopy is aimed at determining the difference in the chemical structure of the reprocessed samples and the witness samples, the protocol characterizes only of the filter media (Melt-blown), by examining changes in the molecular structure. The spectrograms characterizing the chemical structure of the tested samples are presented below (Figure II).

**Figure II.**
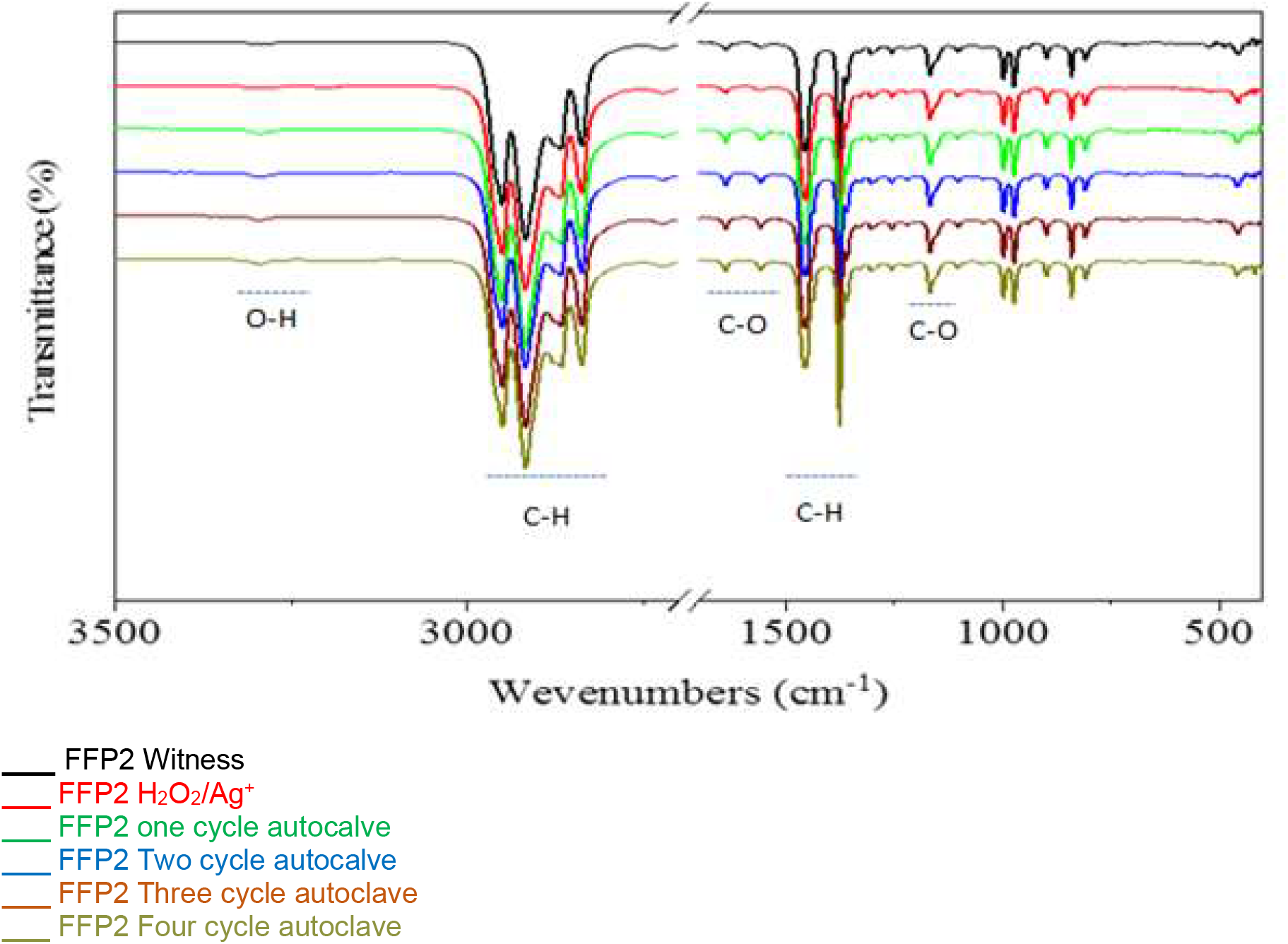
FTIR Spectra of the Melt-blown layers *composing FFP2 masks per reprocessing cycle*

All the dominant bands observed are the characteristic vibration bands of isotactic (iPP) ^26 27^. The FTIR spectrum of our samples shows a strong alkane C-H stretching vibration band from 2841 to 2955 cm^-1^, and the peak at 1380 and 1454 cm^-1^ is represented by the C-H bending.

The emergence of a band around 1725 cm-1 was observed for all samples, the presence of this signal for the spectra indicates the presence of a carbonyl group (C = O) which is derived from the polyester group ^27^. Its presence suggests that in the structure of the analyzed samples there is additionally a PET thermoplastic polymer. The tests confirmed the use of a PP and PET polymer mixture in the Melt-blown layers of the FFP2 masks as protection against harmful bio-aerosol particles ^26^.

The spectra of the samples shown in Figure 2 were similar; no new peaks were detected, meaning that there were no reactions or material interactions when reprocessing our FFP2 masks compared to the control samples.

### Thermogravimetric Analysis (TGA)

The objective of the TGA is to observe possible moisture saturation in the FFP2 layers, especially as we are using a heat reprocessing method as described above. This parameter can influence, the performance of the filtering media, and can also cause bacterial proliferation during storage in an unfavorable environment ^28 10^. This thermogravimetric analysis can also demonstrate a possible cross-linking or decomposition reaction of the polypropylene at low temperature and subsequently a degradation of the FFP2 filtering capacity toward a moist heat reprocessing technique.

The TGA curves of the FFP2 H2O2/Ag+, FFP2 one Cycle Steam autoclave, and FFP2 three Cycle Steam autoclave showed degradation at 315°C versus the samples of FFP2 Witness, FFP2 Four Cycle Steam autoclave, and FFP2 Two-Cycle Steam autoclave showed degradation at 360°C.

The influence of water saturation on our samples retreated with moist heat reprocessing was observed for the FFP2 masks that were recycled two to four times; the degradation was lengthened in time compared to the FFP2 Witness.

The degradation temperature of FFP2 H2O2/Ag^+^ is higher (320°C) compared to FFP2 Witness even though polypropylene is resistant to hydrogen peroxide at room temperature^29^.

The weight loss process of our samples is attributed to the depolymerization of the PP framework ^30^. In general, the results of the Thermogravimetric Analysis confirm that our FFP2 masks give certain stability of the filter layers with a moist heat reprocessing with a sterilization plate not exceeding 125°C, at this temperature value, our TGA curves confirm this stability in figure III.

**Figure III.**
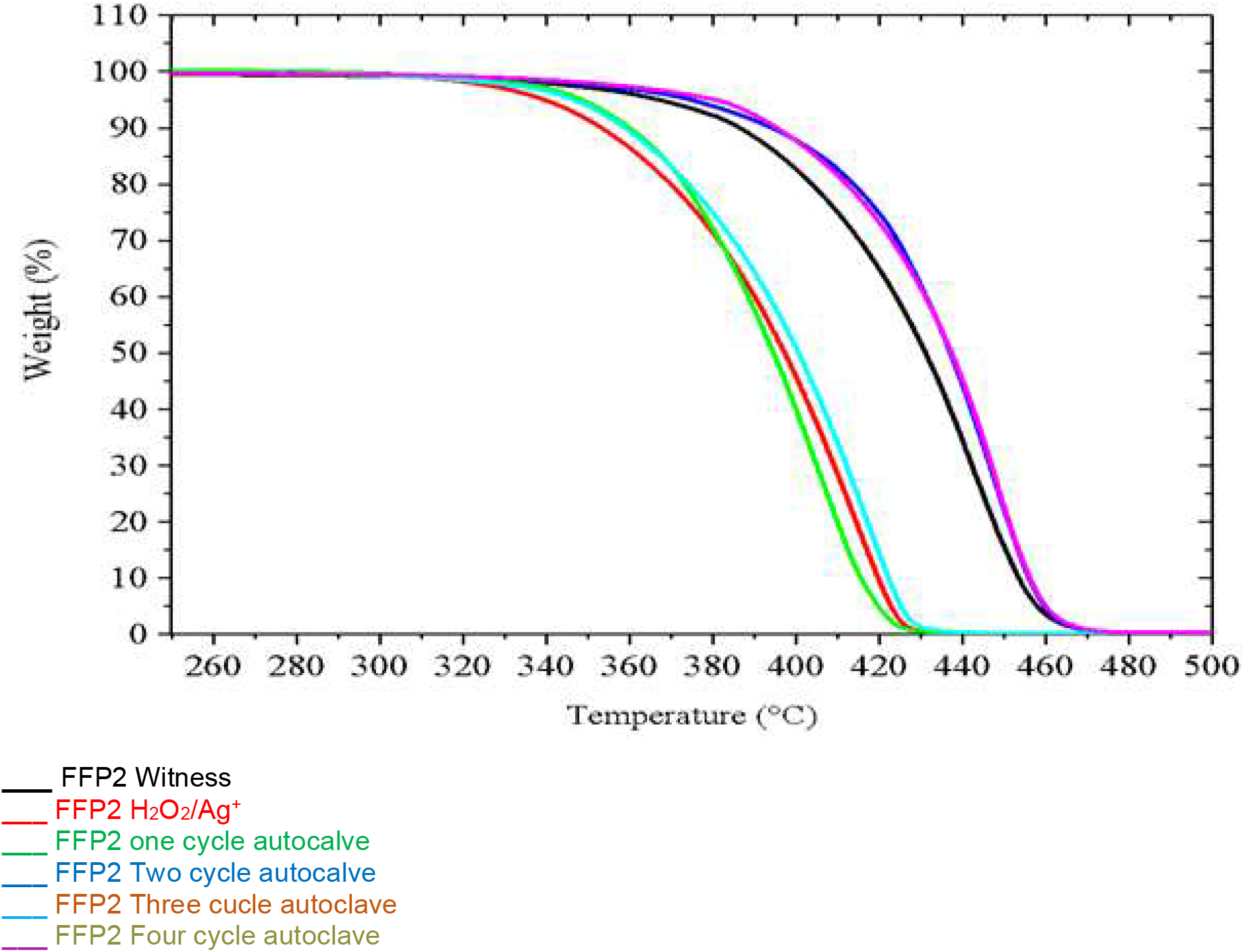
thermogravimetric analysis curves of the layers composing FFP2 masks per reprocessing cycle.

This result indicates that the reprocessing methods do not induce measurable chemical derivative products resulting to cross-linking or chain splitting of polypropylene.

## Discussion

In the rules of good practice for personal protection in healthcare settings, and more specifically respiratory protection, FFP2 masks and other categories of masks have always been recommended for single use. However, the shortage in the supply of PPE in this pandemic context and the duty to preserve a safe working environment for healthcare professionals have emphasized the need to explore creative solutions for PPE shortages.

The proposed reprocessing protocols for FFP2 masks or other respiratory protection still poses problems and technical difficulties related to both, logistical conditions and risk management limitations in the collection and reprocessing chain ^28^. Based on literature data and existing resources, two protocols for reprocessing FFP2 masks have been adopted in this study. One involves the use of hydrogen peroxide doped with silver ions and the other uses moist heat, described as a widely recognized technique in hospital Sterile Processing Systems ^28^.

The undertaken work assessed the feasibility of the reprocessing and the reusing of FFP2 masks, for managing a possible shortage. The results obtained regarding the first reprocessing, based on hydrogen peroxide and silver ions, lead us to conclude that there was no influence on the performance of the tested masks. The characterization tests come to justify the chemical and structural stability of our samples. The presence of silver ions at low doses seemed negligible to cause an effect on the electrostatic filtration provided by the electric charge of the melt (“electret”) used in this type of masks. Another study using the same reprocessing method concluded that the mechanical integrity and performance of the FFP2 is maintained after exposure to 10 or 20 cycles of hydrogen peroxide ^5^.

The concern of inactivation of Sars-CoV has been widely documented; indeed, there is a multitude of scientific research into the inactivation of virus and bacterial spores using hydrogen peroxide ^31 32^. and recently a report submitted to the U.S. FDA by a Columbus company, Battelle, indicates that vaporized hydrogen peroxide (HPV) can be used to safely sterilize N-95 FFR. In its study, the company used spores of the bacterium Geobacillus stearothermophilus, as a biological indicator, to contaminate N-95 FFRs (contaminated either by liquid droplets or by exposure to an aerosol) ^5^.

On the second method of reprocessing the FFP2 masks by moist heat, the results seem to be reassuring for four reuses. Indeed, no modification was noticed regarding the structures or deformation of the integrity of our samples on a sterilization plate at 121°C. A study conducted under the same conditions confirms these results, a permeability test of S. epidermidis and filtration tests, showed no significant difference before and after five cycles of wet heat decontamination ^14 33^. In this regard, another study used moist heat decontamination in real-world testing, reported a moderate influence on the integration of the tested masks at 60°C and 80% relative humidity^34^. while another study indicates that decontamination tests by autoclaving at 134°C in Dutch hospitals resulted in mask deformation and loss of elastic resistance ^35^.

The limitation of this work is that we did not validate the removal of the SARS-CoV-2 virus by autoclaving, but instead relied on multi-parameter chemical indicators to demonstrate the performance of the sterilization load on already biological valid autoclaves. However, a recent study indicated that a sterilization cycle such as ours (15 minutes at 121°C) removed the SARS-CoV-2 ^35^.

## Conclusion

The reuse of respirator masks has been extensively tested during this pandemic period. Various procedures have been proposed and given the variability of the results, likely the success of the decontamination is very closely associated with the mask models and their intrinsic structure.

This work has used more comprehensive testing, by involving tests that aim to detect possible degradation of the material and the possible appearance of derivatives that may be harmful to the caregivers. We have shown in addition to the main objective of the study, that there are no chemical results from the treatment of FFP2 masks under such conditions.

Following our results the proposed reprocessing techniques did not influence the functionality of the tested Duckbill FFP2 masks., in case of an acute shortage of FFP2 masks, these simple processes, hospitals available, economical, and fast to perform, can ensure safe reuse of tested FPP2 masks.

## Data Availability

no data available

## Declaration of conflicting interests

The author(s) declared no potential conflicts of interest concerning the research, authorship, and/or publication of this article.

## Acknowledgments

We are grateful to all who participated in the success of this work from UEMF, CIF, and USMBA of Fez, Morocco, which demonstrates a high level of responsiveness during the SARS-CoV-2 pandemic. We also thank the volunteers who participated in this project

## Funding

This research has not received any specific grants from funding agencies in the public, commercial, or not-for-profit sectors.

## Notes

### Competing Interest Statement

The authors have declared no competing interest.

### Author Declarations

Ethics Committee of the University Hospital of Fez

